# Emerging high-level ciprofloxacin-resistant *Salmonella enterica* serovar Typhi haplotype H58 in travelers returning from India to the Republic of Korea

**DOI:** 10.1101/2020.03.17.20033985

**Authors:** Eunkyung Shin, Ae Kyung Park, Jungsun Park, Soojin Kim, Hyun Ju Jeong, Kyoungin Na, Hyerim Lee, Jeong-hoon Chun, Kyu Jam Hwang, Chul-Joong Kim, Junyoung Kim

## Abstract

In Korea, typhoid fever is a rare disease due to improved living standards. However, this disease remains a major burden in developing countries and regions, such as India and Southeast Asia, In this study, we isolated *Salmonella* Typhi (*S*. Typhi) from eight typhoid patients, travelers returning from India. Isolated strains characterized with antimicrobial susceptibility profile and whole genome sequencing (WGS) analysis.

All strains were resistant to nalidixic acid and azithromycin. Among them, four isolates were highly resistant to ciprofloxacin (MIC ≥32 μg/ml) and never confirmed in Korea *PulseNet* DB. On the basis of the WGS, we found that the ciprofloxacin-resistant strain belonged to global dominant multi-drug resistant (MDR) genotype, haplotype H58 (SNP *glpA* C1047T, *SptP* protein Q185* (premature stop codon)) not harbour MDR plasmid. They have H58-associated SNPs in membranes and metabolism genes, including *yhdA, yajI, hyaE, tryE, rlpB* and *metH*. Also, Phylogenetic analysis showed that H58 strains were assigned sublineage II, and the non-H58 strains were closely related to haplotype H50.

High-level ciprofloxacin-resistant *S*. Typhi haplotype H58 in Korea was first confirmed as influx from overseas by travelers. This study provides information about intercontinental drug-resistant transmission between countries and suggests that travelers need to be careful about personal hygiene.

**Author summary:** Typhoid fever is a systemic human disease including gastroenteritis, fever, and severe diarrhea, caused by *Salmonella enterica* serotype typhi (*S*. Typhi), and requires prompt antibiotic treatment. Due to improved living standards, it has become a rare disease but still prevalent in developing countries such as India and Southeast Asia. Most of the reported case were related with travelers or immigrants from these regions. There is global health problem that emergence of antimicrobial resistant strain associated with fluoroquinolones and third-generation cephalosporins because of leading to treatment failure, serious morbidity, and as well as economic loss. Of antimicrobial resistant *S*. Typhi, haplotype H58 is a dominant multi-drug resistant lineage and has reported in endemic region over past two decades. We identified fluoroquinolones resistance in *S*. Typhi infected after travel to India. Among them, some strains highly resistant to ciprofloxacin were confirmed to have characteristics of haplotype H58. In Korea, *S*. Typhi haplotype H58 from traveller has not been confirmed before. This study provides information about intercontinental drug-resistant transmission between countries and suggests that travelers need to be careful about personal hygiene.

## Introduction

Typhoid fever, caused by *Salmonella enterica* serovar Typhi (*S*. Typhi), is a systemic disease that causes gastroenteritis, fever, and severe diarrhea. *S*. Typhi is a human-specific pathogen transmitted by the ingestion of contaminated water or food(1). This disease remains a major burden in developing countries, such as India, Pakistan, and Egypt, and causes 216,000 deaths/year worldwide(2).

Multidrug-resistant (MDR) *S*. Typhi displaying resistance to chloramphenicol, ampicillin, and trimethoprim has been reported in epidemic countries since the 1960s. Resistance to fluoroquinolones and third-generation cephalosporins was recently demonstrated in the 2000s, and more than 90% of south Asian *S*. Typhi strains have decreased susceptibility to ciprofloxacin (CIP)(3). SNP-based analysis schemes have been developed by stratifying the *S*. Typhi population by haplotype(4). According to Wong et al., over the past two decades, a dominant MDR lineage, *S*. Typhi haplotype H58, initially defined by the SNP *glpA*-C1047T, has become prevalent worldwide. In particular, decreased susceptibility of *S*. Typhi to fluoroquinolones, which is frequently reported in Africa and Asia(5,6), is associated with the H58 sublineage.

Typhoid fever generally occurs in only individuals who have travelled to endemic countries and immigrants from those regions. In Switzerland, quinolone-resistant typhoid infection was reported in a traveler returning from India(7), and MDR *S*. Typhi isolates related to travel to Pakistan were detected in the United Kingdom(8). Typhoid fever is uncommon in the Republic of Korea; approximately 156 cases are reported yearly to the Korea National Surveillance System(9). In 2017, we first isolated H58 *S*. Typhi during an outbreak involving international travelers returning from India. To investigate the transmission route and characterize the isolates, we analysed the H58 strain using whole-genome sequencing.

## Materials and Methods

### Clinical isolates

Stool and blood samples of patients were processed according to the standard bacterial culture method; bacterial identification was performed using a VITEK-II automated system (bioMérieux, Marcy l’Etoile, France), and serotyping of *Salmonella* was carried out according to the Kauffmann-White scheme using antisera (BD Biosciences, Frankland Lakes, NJ, USA). The clinical samples were collected anonymously by *EnterNet* Korea, National gastrointestinal infections disease surveillance network.

### Antimicrobial resistance profile

Antimicrobial susceptibilities and minimum inhibitory concentrations (MICs) were determined using the broth microdilution method with 16 antibiotics and customized KRCDC1F Sensititre panels (Trek Diagnostic Systems, OH, USA) in accordance with the guidelines established by the Clinical and Laboratory Standards Institute (CLSI). The antimicrobial agents tested were ampicillin, ampicillin–sulbactam, amoxicillin–clavulanic acid, cephalothin, cefoxitin, ceftriaxone, cefotaxime, nalidixic acid, ciprofloxacin, tetracycline, chloramphenicol, imipenem, streptomycin, gentamicin, amikacin, and trimethoprim–sulfamethoxazole.

### Whole-genome sequencing

Genomic DNA was isolated with DNA Blood and Tissue Kit (Qiagen, Hombrechtikon, Switzerland). Whole-genome sequencing was performed on a MiSeq platform (Illumina Inc., San Diego, CA) with a v.2 (500 cycles with 2 X 250-nt reads) kit. The existence and mutations of antibiotic resistance genes and plasmids were determined using Bacterial Analysis Pipeline version 1.0.4. (Illumina BaseSpace Labs, https://basespace.illumina.com/). Other alleles in virulence-and adaptation-related genes were identified using the Geneious Prime 2019.2.1 program.

### wgSNP and wgMLST analysis

The paired-end reads were performed with the wgSNP module of BioNumerics v7.6 software (Applied Maths) and mapped to *S*. Typhi reference genome CT18 (AL513382). By choosing the default strict filtering option with minimum coverage x30 base quality, strain-specific SNPs were determined. A maximum likelihood phylogeny was inferred from the identified SNPs using MEGA-X with 1,000 generalized bootstrap replicates, a time-reversible model, and a gamma distribution with an invariant site. The tree was visualized by iTOL. Then, wgMLST was analysed using 15874 loci with the open data set in the wgMLST module. The analysis scheme was developed by Applied Maths, and the default parameters were used. By advanced cluster analysis, a minimum spanning tree was constructed.

## Results

### Case description

In July and August 2017, eight patients were found to be infected with typhoid fever. Five patients had volunteered and travelled in the Northwest region of India (New Delhi, Amritsar, Dharamshala, and Agra) on July 19-27th (group A). The first case occurred on July 22nd and was accompanied by fever, chills, watery diarrhea, headache, and sweating. The onset of the second case occurred on July 28th. After returning to Korea, three patients showed symptoms such as fever, chills, anorexia and stomach-ache on the 6th (n=2) and 13th (n=1) of August. In August, another group travelled along the same route (group B). Three tourists were reported as having typhoid fever on August 18th (Figure 1).

**Figure 1.**
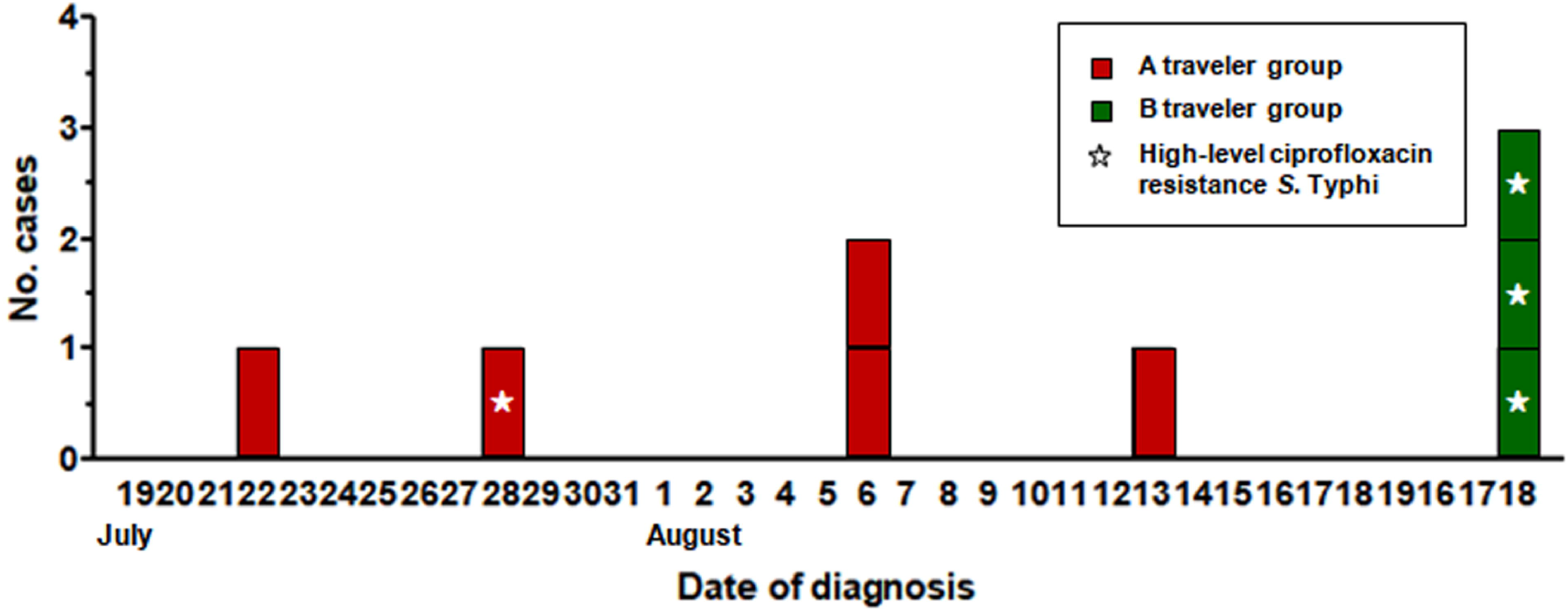
Epidemic curve of typhoid fever outbreak by date of onset. Traveler group A cases are shown in red, and traveler group B cases are shown in green. Cases of high-level ciprofloxacin-resistant *S*. typhi are marked with asterisks.

### Antibiotic resistance profiles

*S*. Typhi was isolated from all eight patients and determined by the antimicrobial susceptibility test. All strains were resistant to nalidixic acid (NAL; minimum inhibitory concentration (MIC) ≥ 128 μg/ml) and azithromycin (AZI; MIC ≥ 8 μg/ml). Only four isolates (1 from group A and 3 from group B) were highly resistant to ciprofloxacin (MIC ≥ 32 μg/ml) (Table 1). Furthermore, analysis of the antimicrobial gene mutations of the CIP-resistant isolates revealed two mutations in the *gyrA* gene (Ser83→Phe (S83F) and Asp87→Asn) and a *parC* mutation (Ser80→Ile). In addition, the analysis of plasmid-mediated quinolone resistance (PMQR) determinants indicated that the CIP-resistant isolates contained the *aac(6’)-Iaa* gene. The non-H58 strain had a mutation in *gyrA* (S83F) and a PMQR gene (*aac(6’)-Iaa*) on the plasmid and replicon type (IncFIB (pHCM2)). Pulsed-field gel electrophoresis (PFGE) results indicated that the four CIP-resistant isolates had identical pattern types, while the four CIP-susceptible isolates had another identical type (Appendix figure 1). Notably, neither of the above mentioned PFGE pattern types has been assigned before in *PulseNet* Korea

**Table 1.**
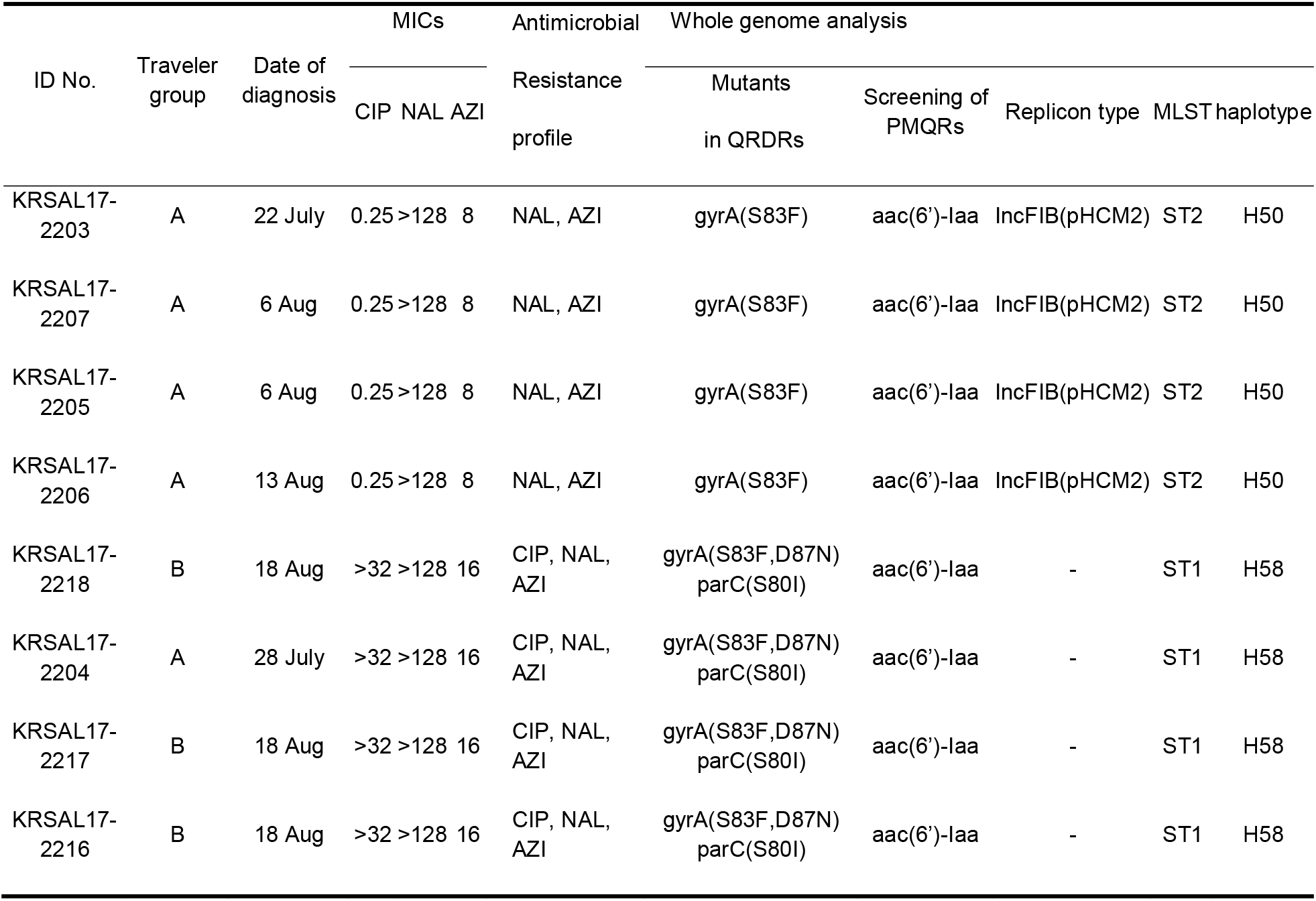
Antimicrobial resistance profiles and genomic analysis of the outbreak strains.

### Whole-genome sequencing analysis

whole-genome sequencing (WGS) analysis indicated that high-level CIP-resistant strains belonged to haplotype H58, while the CIP-susceptible strains did not. The CIP-resistant strains had the typical features of H58: SNPs in *glpA* at nucleotide 252 and position 2,348,902 in *S*. Typhi CT18 (C1047T) and *SptP* protein Q185* (premature stop codon). Maximum likelihood phylogeny was inferred from 1577 SNPs (Figure 2A). SNP-based analysis distinguished the *S*. Typhi isolates by haplotype. The H58 strains formed a tight cluster and belonged to sub-lineage II, with comparative strains from India, Viet nam, Sri lanka and Lebanon. They have H58-associated SNPs in membranes and metabolism genes, including *yhdA, yajI, hyaE, tryE, rlpB* and *metH*. The non-H58 strains were closely related to haplotype H50. A minimum spanning tree (Figure 2B) based on wgMLST showed similar results, but it is difficult to distinguish between H58 sub-lineages.

**Figure 2.**
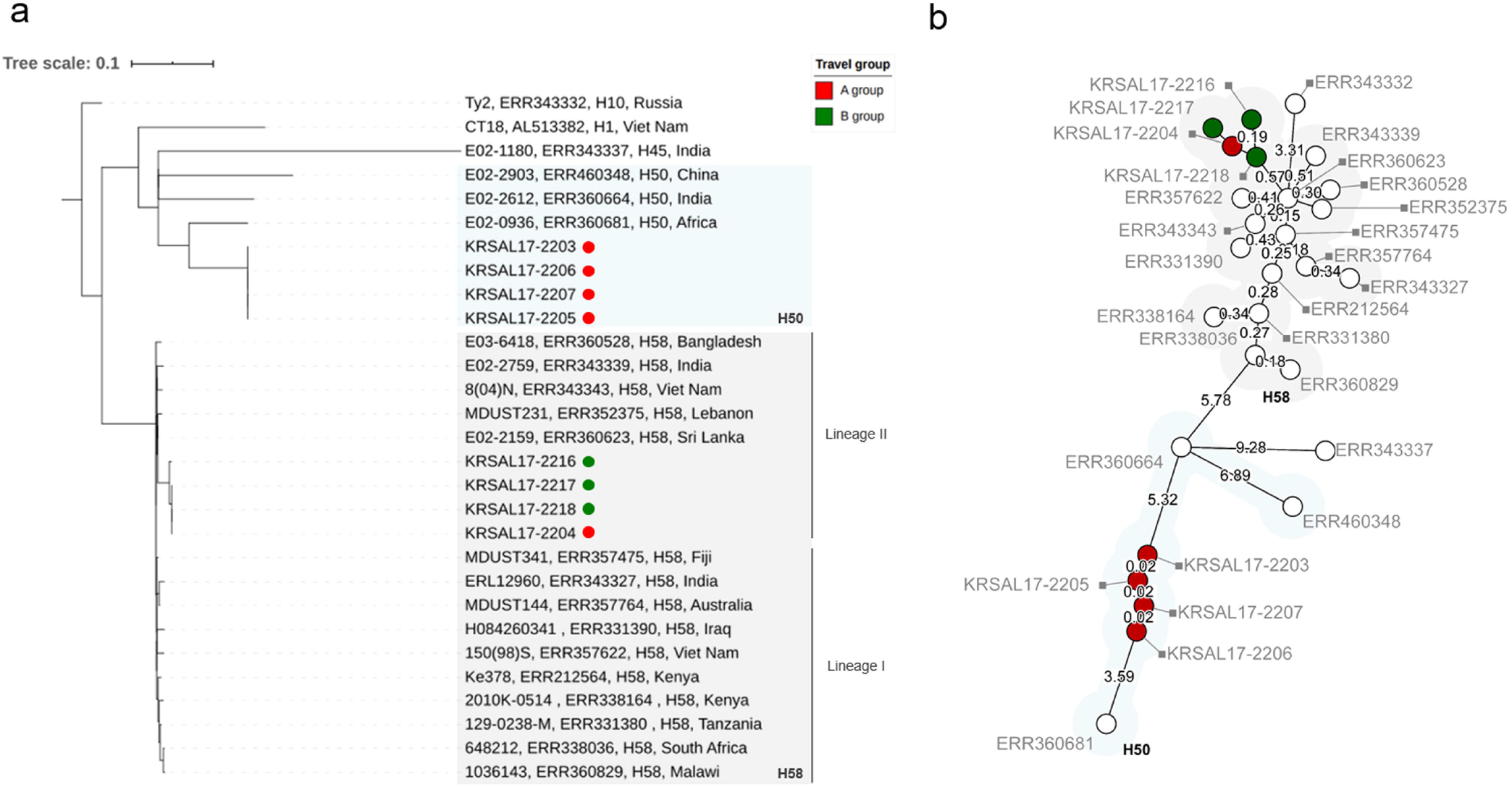
Whole-genome sequencing trees of *S*. Typhi isolates based on SNP and wgMLST analyses. A, Maximum likelihood tree inferred from the 1577 identified SNPs and visualized by iTOL. Comparative strains from the EBI (European Bioinformatics Institute) database and isolates are marked in the following order: isolate name, Short Read Archive (SRA) accession number, haplotype, and country. The numbers above the nodes represent data from bootstrap replicates. B, Minimum spanning tree based on numerical values (15874 wgMLST loci). The branch lengths are represented on a logarithmic scale. Traveler group A cases are shown by red circles, and traveler group B cases are shown by green circles. The cluster of H58 strains is highlighted in light grey, and the H50 strains are highlighted in light blue.

## Discussion

In Korea, typhoid fever is rare due to improved living standards. Six travel-associated cases (in addition to this outbreak) and 304 *S*. Typhi isolates have been sporadically identified over the past decade, (10). The previous travel cases were associated with China (n=2), Japan (n=1), Indonesia (n=2), and Russia (n=1). India is a well-known geographical source of H58 with antimicrobial resistance (11).). But still, there are no any cases related epidemic region. Therefore, this report describes initial influx of *S*. Typhi H58 outbreak following international travel.

Since the 1960s, antimicrobial-resistant *S*. Typhi has been prevalent worldwide. NAL-resistant *S*. Typhi is frequently isolated in India, and high-level CIP-resistant *S*. Typhi was recently isolated from a domestic patient(12). In 2007, NAL-resistant *S*. Typhi strains were reported for the first time in Korea and did not respond to CIP as treatment(13). These cases suggest an alternative treatment as third-generation cephalosporins or AZI (6). But the isolates showed resistance to not only NAL and CIP but also AZI. Moreover, the occurrence with extended spectrum β-lactamases (ESBLs) harbouring CTX-M-15 and/or high resistance to quinolones has been reported in other studies(14). Therefore, an appropriate prescription based on surveillance will be required for future treatment. Continuous monitoring and data collection will be essential to prevent the occurrence and treatment failure of antimicrobial-resistant *S*. Typhi.

As the dominant MDR lineage of *S*. Typhi, haplotype H58 has spread worldwide, particularly in South Asia and Africa (3,4). In this research, we revealed H58 *S*. Typhi infection by international travelers using WGS analysis. These H58 strains harboured a *glpA* gene and *SptP* protein mutation and triple point mutations in QRDR but lacked the MDR plasmid of the IncHI1 type. The most of H58 lineage relate with reduced globally susceptibility to fluoroquinolones and harbouring IncHI1 (4). Our results indicate that fluoroquinolone-resistant H58 with extending other antimicrobial resistance is circulating in epidemic regions. Thus, continuous surveillance will be needed to monitor intercontinental transmission of antimicrobial-resistant strains and support the global management of typhoid fever.

## Data Availability

The whole-genome data of this study have been deposited in BioProject under number PRJNA551786. The raw reads are available in the Sequence Read Archive (SRA) under accession number SRR9616298-SRR9616305.

## Ethical considerations

The Ethics committee of the first affiliated Korea Centers for Disease Control and Prevention decided that Institutional Review Board approval was not required. Because patient information was collected anonymously and confidential patient information was not included.

